# Genetic assessments of breast cancer risk that do not account for polygenic background are incomplete and lead to incorrect preventative strategies

**DOI:** 10.1101/2021.08.13.21262050

**Authors:** George B. Busby, Paul Craig, Nesrine Yousfi, Saurabh Hebbalkar, Paolo Di Domenico, Giordano Bottà

## Abstract

Breast cancer is the most common cancer among women and is a leading cause of cancer mortality worldwide. There is a significant genetic component to breast cancer risk which is the result of both rare pathogenic mutations and common genome-wide variation. However, the penetrance of pathogenic mutations varies widely and their frequency is low, both at a population level and amongst breast cancer cases. Polygenic risk scores, which aggregate the effect of hundreds to millions of common genome-wide variants offer a way to further understand the contribution of genetics to disease risk. Here we analyse genome-wide data from 221,479 women and 90,307 high coverage exomes to understand how rare and common variation affect lifetime breast cancer risk. We show that PRS strongly modulates the penetrance of mutations in 8 breast cancer susceptibility genes. For example, lifetime risk in *BRCA1* carriers with low polygenic risk is almost one third that of carriers with high PRS (26% v 69% in the bottom and top PRS deciles, respectively). Adding family history of breast cancer provides additional stratification on the potential outcome of disease in carriers of rare mutations. PRS also identifies a significant fraction of the population at equivalent risk to carriers of moderate impact pathogenic variants and who are an order of magnitude more common at a population level. These results have important implications for breast cancer risk mitigation strategies, indicating that the genetic risk of breast cancer is determined by both monogenic mutation and polygenic background, and that assessments of genetic risk for breast cancer risk that do not consider the polygenic background are imprecise and unreliable.

## Main Text

Genetic testing for the presence of pathogenic mutations is an established medical practice^1^. However, whilst screening can identify women who are genetically susceptible to breast cancer (BC), two central challenges remain. The first is that pathogenic mutations are rarely completely penetrant, so the presence of a mutation does not always lead to disease^2^. Even for carriers of mutations in the BC susceptibility genes with the highest risk, penetrance ranges from 47% to 66% for *BRCA1* and 40% to 57% for *BRCA2*^3^, meaning that around a third to a half of the women who carry a *BRCA1/2* mutation will not go on to develop BC. For moderate risk BC susceptibility genes such as *PALB2*, penetrance estimates are lower, between 26% and 46%^4^, and carriers of mutations in low risk BC susceptibility genes such as *CHEK2* and *ATM* have a roughly two fold increased risk compared to non-carriers^5,6^. Family history of BC also increases risk by around two times in the absence of other risk factors and further increases penetrance in pathogenic mutation carriers^7,8^. Understanding the additional factors that contribute to modulating the penetrance of variants in these genes is critical to evaluating their clinical utility.

The second challenge is that pathogenic mutations in BC susceptibility genes are rare, both at a population level and among cases. In general, the frequency of a pathogenic mutation is negatively correlated with the risk it confers, so those variants that increase risk the most - and are therefore more penetrant - are rarer in the population^9^. Estimates of mutation prevalence in 12 major BC susceptibility genes from population-based analyses, which are subject to less selection bias than analyses of pedigrees, are around 5% across all 12 genes in BC cases and 1.5% in non-cases^9–12^. At an individual gene level, frequencies range from around 0.11% in *BRCA1* and *PALB2*, 0.25% in *BRCA2*, to 0.41% in *ATM* and *CHEK2* ^11^. Therefore, screening for rare pathogenic mutations will only ever identify a small subset of those at heightened genetic risk of BC.

To address these critical challenges we analysed data from the UK Biobank to ask two questions. First, does incorporating polygenic background to lifetime risk assessments for BC in mutation carriers help explain the incomplete penetrance of these mutations. Second, does polygenic risk provide an additional source of genetic risk that can be used to identify women at high risk of BC in the absence of rare pathogenic mutations.

We began by building a new Polygenic Risk Score (PRS) for BC using Training data (summary statistics) from a large Genome-Wide Association Study of BC^13,14^, together with separate and independent PRS Validation (N=72,421; 4,777 BC cases) and PRS Testing (N=160,469; 10,915 BC cases) datasets of self-identified White British individuals from the UK Biobank^15^ and phenotypes defined in Supplementary Table 1. We applied Allelica’s DISCOVER module^14^ that assesses the performance of 10 different PRS algorithms resulting in a best performing panel that used the Stacked Clumping and Thresholding algorithm^16^. The resulting panel (Allelica 577k) comprised 577,113 genome-wide variants and had increased predictive performance compared with a commonly used 313 variant BC PRS^17^ on the same PRS Testing dataset (Area Under the Receiver Operator Curve (AUC): Allelica 577k = 0.71 (95%CI 0.698-0.717); Mavaddat 313 = 0.68 (95% CI 0.669-0.688); Odds Ratio per standard deviation: Allelica 577k = 1.81 (95%CI 1.78-1.84); Mavaddat 313 = 1.56 (95%CI 1.53-1.58)).

Next, we used whole exome sequence data to identify carriers of likely pathogenic / pathogenic variants (LP/P) according to ClinVar^18^ in 90,307 self-identified British ancestry women in the UK Biobank^19^ (Supplementary Tables 2 and 3). After removing individuals with prevalent BC (N=6,322), we constructed a separate Cox proportional hazards model for each of 8 BC susceptibility genes using BC case status, age of enrollment and age of disease in the survival function, and carrier status, PRS, family history, 4 principal components of ancestry and genotyping array as covariates. For comparison, we built an equivalent model using all women in the PRS Testing dataset that did not include carrier status. We used these models to predict lifetime risk for each percentile of the PRS distribution (Figure 1).

**Figure 1:**
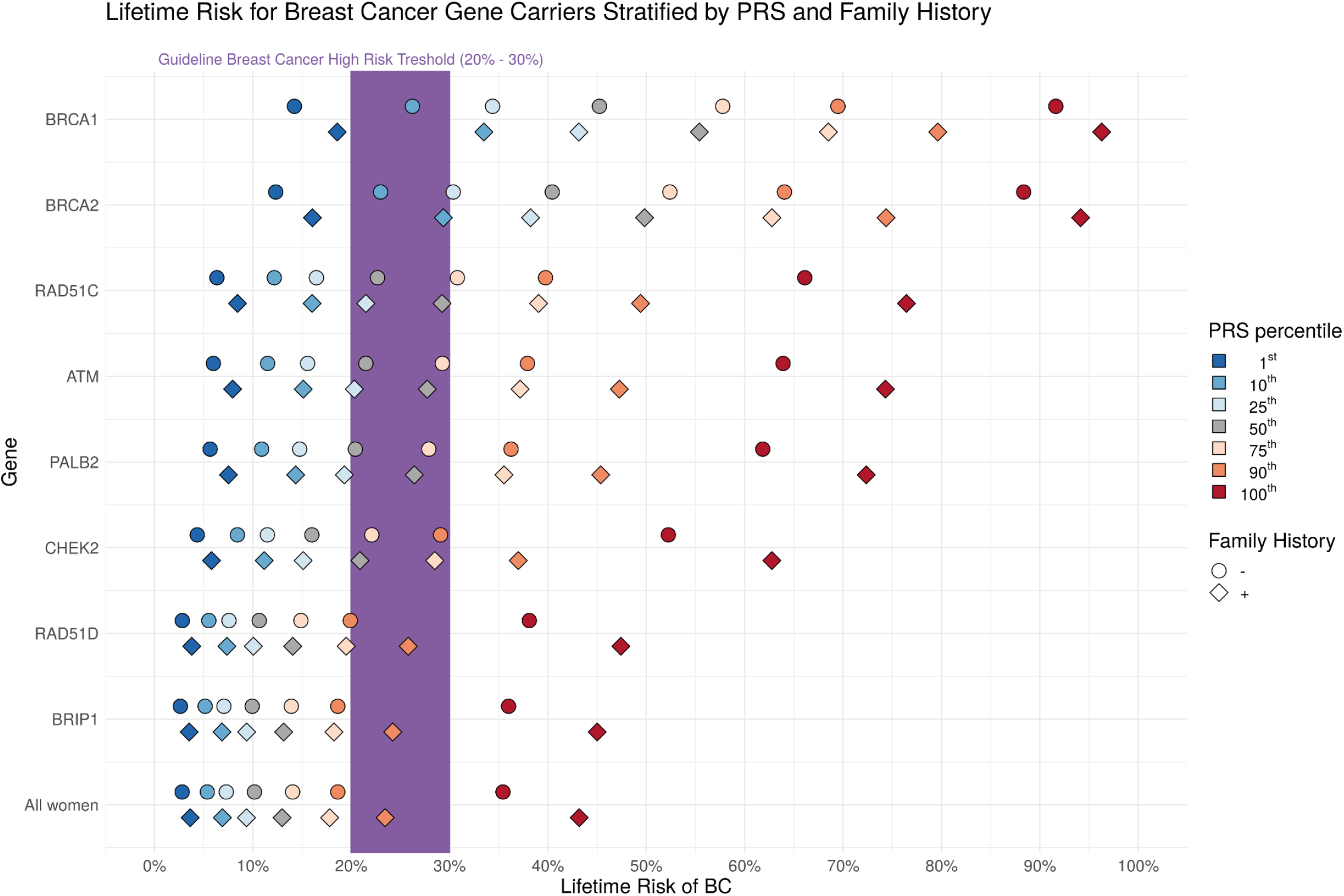
PRS modulates the penetrance of 8 BC susceptibility genes. We identified carriers of pathogenic variants in 8 breast cancer susceptibility genes in whole exome sequence data in 86,385 women in the UK Biobank. For each gene we show the predicted lifetime risk for carriers with (diamonds) and without (circles) family history across a range of different PRS percentiles using the Allelica 577k PRS. Data and confidence intervals are reported in Supplementary Tables 4 and 5. The same analysis using the Mavaddat 313 SNP PRS ^17^ is shown in the Supplementary Figure. An equivalent model without carrier status was built using All women in the PRS Testing dataset. Dashed lines show lifetime risk for percentiles of the Allelica 577k PRS distribution in Non-carriers. American Cancer Association guidelines state that a threshold of 20% lifetime risk equates to High Risk ^20^. We estimated lifetime risk as risk of BC by age 79 with a Cox proportional hazards regression for each gene separately in a model that included case/control status, age of enrollment and age of disease in the survival function, and carrier status, PRS, 4 PCs and genotyping array as covariates.

Amongst carriers of LP/P variants, PRS stratified lifetime risk, extending recent studies demonstrating the modulation of penetrance by BC PRS^21,22^ to a larger set of genes on a larger dataset. Approximately 11% (N=9,669) of the women in this dataset had family history of BC (defined as at least one first degree relative diagnosed with BC) allowing for the additional stratification of risk in carriers with family history to be assessed. Lifetime risk of carriers of mutations in all 8 genes varied substantially on the basis of PRS (Figure 1). Family history provides additional information that modulates BC risk in combination with PRS. Whilst there is no universal definition of high risk for BC, the American Cancer Society suggests a threshold for high risk of 20-25% lifetime risk^20^ and in the US lifetime risk of greater than 20% can lead to the initiation of screening with magnetic resonance imaging (MRI)^23^. Guidelines from the National Institute for Health and Care Excellence in the United Kingdom use two thresholds of risk: Moderate lifetime risk 17-30%; and High lifetime risk >30%^24^. Here we use a broad threshold of 20-30% as High risk. Carriers of LP/P mutations in *BRCA1* in the lowest decile of the PRS distribution without family history have lifetime risk within this High risk threshold (26%; 95% CI: 9-43%), whereas those in the bottom decile with family history have a 1 in 3 lifetime risk of disease (33% (95%CI 13%-54%). At the other end of the distribution, the combination of high polygenic and monogenic risk leads to 80% (95%CI: 55-100%)) and 69% (95% CI: 43-96%) lifetime risk for women in the top decile of the PRS distribution, with and without family history respectively. Around half of LP/P mutation carriers in moderate penetrance genes (*RAD51C, ATM* and *PALB2*) will have an overall lifetime risk of less than 25%. Conversely, women who carry mutations in these genes and in the top percentile of the PRS distribution have a greater than one in two lifetime risk of BC (Figure 1, Supplementary Tables 3 and 4). The effect of PRS on lifetime risk in carriers of variants in *CHEK2, RAD51D* and *BRIP1* is modulated in a similar manner to non-carriers. Taken together, these results indicate that PRS provides additional information on the penetrance of mutations in BC susceptibility genes that can be used by patients and physicians to inform potential risk mitigation options.

We compiled a list of published Odds Ratios (ORs) for 8 BC susceptibility genes, together with their estimated prevalence and compared them with ORs for different strata of the Allelica 577k PRS distribution (Figure 2, Supplementary Table 5)^11,12^. ORs for *BRCA1* carriers are the highest, ranging from point estimates of 7.62 to 10.57. Odds Ratios for the top 0.5% of the Allelica 577k PRS distribution, *BRCA2* and *PALB2* mutation carriers are broadly similar (Figure 2) despite the prevalence of this PRS strata being double that of *BRCA2* carriers (prevalence: 0.24-0.26%) and over four times greater than *PALB2* mutation carriers (prevalence: 0.10-0.12%). PRS therefore identifies a greater number of women at high risk (5X increased risk) than carriers of variants in these genes.

**Figure 2:**
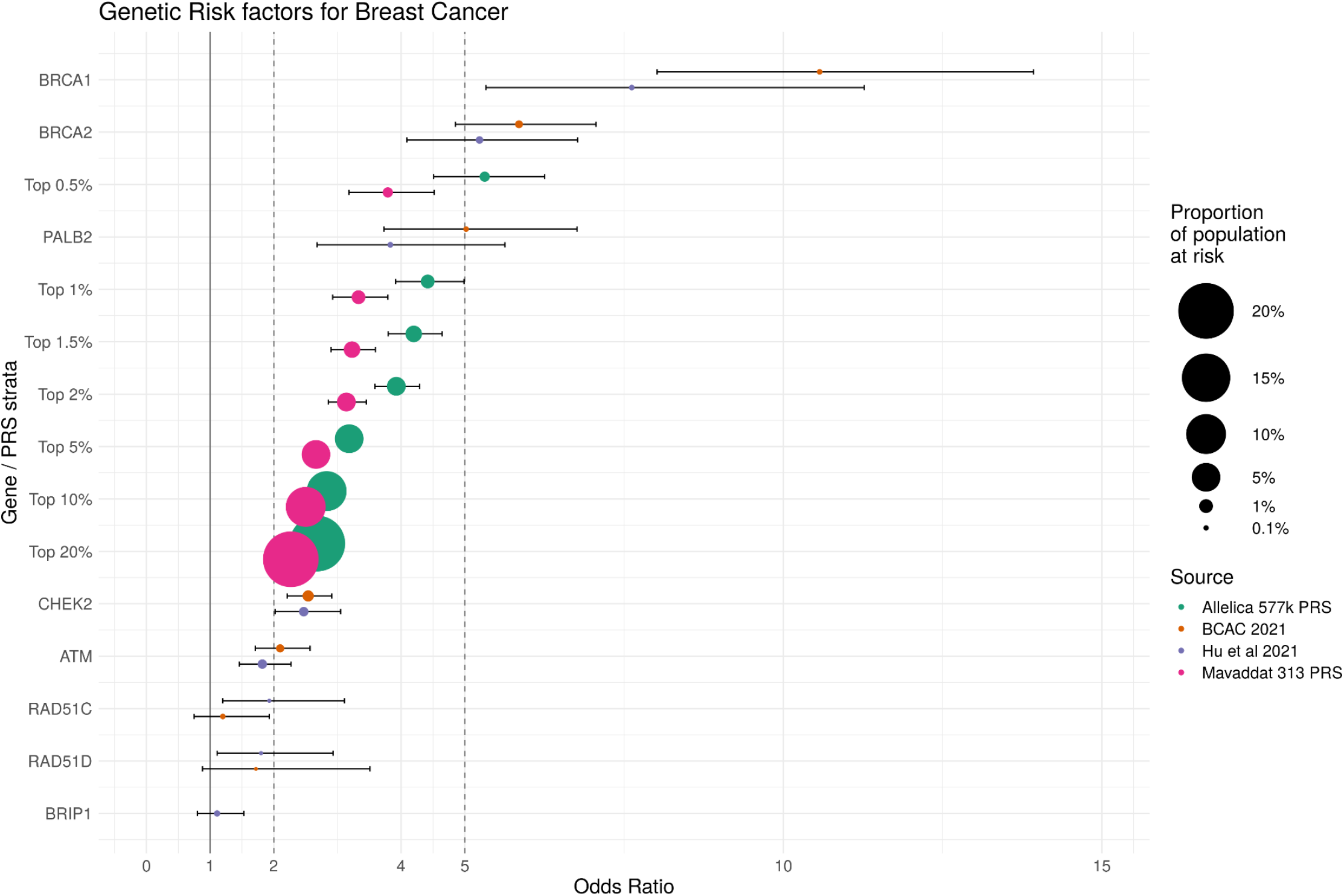
Genetic risk factors for breast cancer. For eight Breast Cancer susceptibility genes we show published estimates of ORs from two separate population-based studies (with the exception of *BRIP1* in the BCAC 2021 study, which was not analysed). For comparison we show ORs for the top X% for the PRS distribution for the Allelica 577k and Mavaddat 313 PRS, which were computed with logistic regression comparing individuals with a PRS in or above the percentile against the remainder of the PRS distribution controlling for the first four principal components of genetic ancestry and age of enrollment. The point size represents the proportion of the population at risk and reflects the number of individuals that have the genetic risk factor.

Genetic factors that increase risk by 2-4 times are considered to be moderate-risk^9^. PRS can identify one in five women (top 20%) with an average genetic risk greater than 2 (OR= 2.68 (95%CI 2.57-2.78)), which is above that for carriers of mutations in *ATM* and equivalent to *CHEK2* mutation carriers. By definition this strata contains 20% of the population, demonstrating the potential for PRS to identify women at both moderate and high genetic risk of BC, who are up to 40 times more common in the population. Risk for women in the top 2% of the PRS distribution approaches four times (3.92 (95%CI 3.58-4.29)) the average population risk. In addition to the Allelica 577k PRS, we also show ORs for PRS percentiles using the Mavaddat 313 SNP PRS^17^ (Supplementary Figure, Supplementary Table 5), which show a similar trend with less intense stratification.

This study supports the hypothesis that common genetic variation controls the penetrance of pathogenic mutations in *BRCA1* and *BRCA2*^25^ as well as moderate penetrance genes. Together with information on pathogenic carrier status and family history, PRS has the potential to inform current screening approaches^26^. For example, *BRCA1* and *BRCA2* mutation carriers with no family history of BC who are in the lowest decile of the PRS distribution have a lower than 1 in three lifetime risk of BC, information that can be used to guide and potentially avoid invasive risk mitigation strategies involving surgery intervention such as mastectomy. Around half of carriers of mutations in moderate penetrance genes have a less than 20% lifetime risk of BC, whereas those with the highest PRS scores, for example in the top decile of the PRS distribution (Figure 1) have over double the lifetime risk (∼40%). When combined with family history, this risk rises still further. In the absence of pathogenic mutations, PRS can identify 1% of women with a greater than 4 fold increased risk of BC (Figure 2). One strategy where PRS could have immediate utility is as an additional screening tool for women identified as high risk through other risk factors, such as family history^23^. Whilst family history is a robust predictor of breast cancer risk, one striking outcome of this present study is that women with rare pathogenic mutations and family history can have wildly different risk trajectories because of their polygenic risk. For example, lifetime risk for women with family history of BC who are carriers of a pathogenic mutation in *BRCA2* varies from less than 30% in the bottom decile to almost 75% in the top decile of the PRS distribution (Supplementary Table 4).

This work has some limitations. First, our analyses are based on a European ancestry subset of the UK Biobank and results may translate differently across ancestries. PRS is not alone in having an ancestry bias: for example, the ability of the commonly used GAIL model to assess 5 year breast cancer varies in different ancestry groups^27^ and pathogenic founder mutations in *BRCA1/2* genes are under-represented in clinical databases^28^, meaning that screening for known variants is less effective in Non-European ancestry groups. As such, current guidelines on the clinical utility of these variants take a woman’s ancestry into account. A similar approach can be taken with PRS, where well validated, ancestry-specific scores can be used when they outperform those developed on predominantly European ancestry datasets. We support all efforts to build ancestrally diverse datasets to ensure that genomic information can be used widely. Secondly, environmental factors contribute to a woman’s risk^29^ which in the current context may include modulating the penetrance of BC mutations and the risk conferred by the PRS. Thirdly, despite utilising data from over 90 thousand exomes, mutation carriers are rare (Supplementary Table 2). Larger datasets will allow for further granularity of the penetrance modulation estimates presented here.

Current guidelines do not include the use of PRS to assess clinical risk of BC. Whilst the benefits of assessing individuals for the presence of pathogenic mutations in BC susceptibility genes has undoubted utility in those that carry them, PRS provides fundamental and complementary information on the effect of these variants on the lifetime risk of disease that can be used to guide preventative strategies according to current guidelines. In addition, PRS identifies women at high genetic risk who are invisible to screening of rare pathogenic mutations. Without considering PRS in pathogenic mutation testing, about half of the carriers of mutations in moderate penetrance genes are incorrectly assigned as high risk, leading towards potentially unnecessary preventive measures. Even with a family history of breast cancer, around 10% of *BRCA1* carriers have a 1 in 3 lifetime risk of BC. Whilst high enough to warrant active surveillance through mammographic screening and MRI, this risk level is not high enough to justify invasive, unnecessary and potentially harmful surgical intervention. For the 95% of women whose BC is not the result of rare pathogenic mutations, PRS offers a way to identify heightened risk, comparable to that conferred by monogenic mutations yet present in significantly more women. Combining information from both rare and common variation with family history paves the way for a targeted and data-driven approach to BC risk mitigation.

## Methods

### Data

This study utilises both the UK Biobank imputed genotype and whole exome sequence data under project number 40692. Full details of the UK Biobank resource have been described previously^15^. UK Biobank received ethical approval from the North West Multi-Center Research Ethics Committee (11/NW/03820). We used data from a subset of women self-identified as having White British ancestry (N=221,479) and further split this dataset to produce independent Validation and Testing datasets using the UK Biobank first release (genotyping batch from 11 to 22) and second release (genotyping batch from 23 to 95), respectively. Breast cancer phenotype information was collated by extracting data in the fields reported in Supplementary Table 1. The Validation dataset comprised 4,764 BC cases (1,811 incident / 2,953 prevalent) and 64,055 controls and the Testing dataset comprised 10,871 cases (4,163 incident / 6,708 prevalent) and 141,789 controls.

### Building a new PRS for Breast Cancer

We applied the following algorithms to identify the PRS with the best predictive performance: Clumping and Thresholding^30^, Stacked Clumping and Thresholding^16^, LDPred2, LDPred2-Inf, LDPred-funct^31^, Lassosum^32^, SBayesR^33^, PRS-CS, PRS-CS-auto^34^ and an ensemble score generated using machine learning (Support Vector Machine). PRS panels were developed using Training data (Genome Wide Association Study summary statistics) from Michaillidou et al^13^, and the separate Validation and Testing datasets outlined above. The score with the best predictive power (highest Area Under the Curve) in the Validation dataset used the Stacked Clumping and Thresholding algorithm and contained 577,113 genome-wide variants. AUC and Odds Ratio per Standard Deviation are reported in the Testing dataset.

### Classifying pathogenic variants

We used the *TAPES* software^35^ to classify all variants in 90,307 women with self-identified White British ancestry from the UK Biobank whole exome sequence dataset. Of these 6,332 were BC cases (3,992 prevalent / 2,410 incident) and 83,975 were controls. All variants identified as Likely Pathogenic / Pathogenic (LP/P) according to ClinVar ^18^ in each of the following 8 breast cancer susceptibility genes were kept for further analysis: *ATM, BRCA1, BRCA2, BRIP1, CHEK2, PALB2, RAD51C* and *RAD51D*.

### Lifetime risk

Lifetime risk of breast cancer was computed with Cox proportional hazards models using the coxph function from the *survival* package in *R*^36^. The survival function contained breast cancer cases status (for incident cases and controls only) together with age of disease, age of enrollment. Standardised PRS, first four ancestry PCs and genotyping arrays were used as covariates together with carrier status where relevant. Lifetime risk was predicted by predicting survival by age 79 for each percentile of the PRS distribution using the *riskRegression* package in *R*^37^. To account for non-normally distributed risk within PRS percentiles, lifetime risk was predicted for each 0.01% of the distribution and then averaged across a given percentile by fitting an exponential curve to the data within a percentile and finding the rate of this curve.

### Association testing

We used the glm function in R to perform logistic regression to calculate odds ratios and 95% confidence intervals for top X% of the PRS distribution. Breast cancer case status was predicted using an independent indicator variable for each individual’s assignment to the top X% of the PRS distribution (where X = 80,90,95,98,99,99.5), and controlling for the first four ancestry PCs, genotyping array, age of enrollment.

## Data Availability

This work uses data from the UK Biobank which is available to researchers via application.

https://www.ukbiobank.ac.uk/

## Acknowledgements

We thank all participants in the UK Biobank. This work was funded by internal research and development funding from Allelica, Inc.

## Competing Interests

All authors are employees at Allelica Inc.

## Supplementary Material

**Supplementary Table 1:** UK Biobank codes used to define breast cancer phenotypes used in this study.

| Breast Cancer (BC) |  |  |
| --- | --- | --- |
| UK BioBank Field | Description | Codes |
| 20001 | Self reported cancer code | 1002 |
| 41202 | Diagnoses -main ICD10 | C500-C509,<br>D050-D051,<br>D057,D059 |
| 41204 | Diagnoses -Secondary ICD10 |  |
| 40001 | Underlying cause of death - ICD10 |  |
| 40002 | Contributory causes of death - ICD10 |  |
| 40006 | Type of cancer - ICD10 |  |

**Supplementary Table 2:** Number of carriers of pathogenic mutations in 86,385 women in the UK Biobank. The number of carriers of likely pathogenic or pathogenic variants (L/LP) according to ClinVar. The number of unique variants in each gene as well as the number of cases who are carriers is also shown. This analysis was conducted on incident BC cases only.

| Gene | Num. carriers | Num. unique variants | Num. cases |
| --- | --- | --- | --- |
| BRCA1 | 46 | 32 | 7 |
| BRCA2 | 210 | 105 | 27 |
| RAD51C | 32 | 14 | 2 |
| ATM | 261 | 101 | 17 |
| PALB2 | 77 | 30 | 5 |
| CHEK2 | 119 | 24 | 5 |
| RAD51D | 35 | 9 | 1 |
| BRIP1 | 104 | 31 | 3 |
| **Total** | **884** | **346** | **67** |

**Supplementary Table 3:** Number of carriers of pathogenic mutations in 3,922 women in the UK Biobank with prevalent breast cancer (i.e. before enrollment in the UK Biobank). The number of carriers of likely pathogenic or pathogenic variants (L/LP) according to ClinVar. The number of unique variants in each gene as well as the number of cases who are carriers is also shown. These individuals were removed from the association analyses in this study.

| Gene | Num. carriers | Num. unique variants | Num. cases |
| --- | --- | --- | --- |
| BRCA1 | 25 | 24 | 25 |
| BRCA2 | 56 | 43 | 56 |
| RAD51C | 1 | 1 | 1 |
| ATM | 36 | 27 | 36 |
| PALB2 | 7 | 7 | 7 |
| CHEK2 | 9 | 4 | 9 |
| RAD51D | 4 | 4 | 4 |
| BRIP1 | 9 | 7 | 9 |
| **Total** | **147** | **117** | **147** |

**Supplementary Table 4:** Predicted Lifetime risk (risk by age 79) and 95% confidence intervals for carriers of pathogenic mutations without family history of breast cancer at different PRS strata using the Allelica 577k and Mavaddat 313.

| Gene | PRS percentile | Lifetime Risk (Allelica 577k) | Lifetime Risk (Mavaddat 313) | Gene | Lifetime Risk (Allelica 577k) | Lifetime Risk (Mavaddat 313) |
| --- | --- | --- | --- | --- | --- | --- |
| BRCA1 | 1 | 0.14 (0.04-0.24) | 0.17 (0.05-0.29) | CHEK2 | 0.04 (0.01-0.08) | 0.05 (0.01-0.09) |
| BRCA1 | 10 | 0.26 (0.09-0.43) | 0.29 (0.1-0.47) | CHEK2 | 0.08 (0.01-0.15) | 0.09 (0.01-0.16) |
| BRCA1 | 25 | 0.34 (0.14-0.55) | 0.36 (0.14-0.58) | CHEK2 | 0.11 (0.02-0.21) | 0.11 (0.02-0.21) |
| BRCA1 | 50 | 0.45 (0.21-0.7) | 0.45 (0.21-0.7) | CHEK2 | 0.16 (0.03-0.29) | 0.15 (0.03-0.28) |
| BRCA1 | 75 | 0.58 (0.31-0.85) | 0.56 (0.29-0.83) | CHEK2 | 0.22 (0.05-0.39) | 0.2 (0.04-0.36) |
| BRCA1 | 90 | 0.69 (0.43-0.96) | 0.66 (0.38-0.93) | CHEK2 | 0.29 (0.08-0.5) | 0.25 (0.06-0.45) |
| BRCA1 | 100 | 0.92 (0.77-1) | 0.87 (0.67-1) | CHEK2 | 0.52 (0.22-0.83) | 0.43 (0.15-0.71) |
| BRCA2 | 1 | 0.12 (0.08-0.17) | 0.15 (0.09-0.21) | RAD51D | 0.03 (0-0.08) | 0.04 (0-0.11) |
| BRCA2 | 10 | 0.23 (0.15-0.31) | 0.25 (0.16-0.34) | RAD51D | 0.06 (0-0.16) | 0.06 (0-0.19) |
| BRCA2 | 25 | 0.3 (0.2-0.4) | 0.32 (0.21-0.42) | RAD51D | 0.08 (0-0.22) | 0.08 (0-0.24) |
| BRCA2 | 50 | 0.4 (0.28-0.53) | 0.41 (0.28-0.53) | RAD51D | 0.11 (0-0.31) | 0.11 (0-0.32) |
| BRCA2 | 75 | 0.52 (0.38-0.66) | 0.51 (0.37-0.64) | RAD51D | 0.15 (0-0.42) | 0.15 (0-0.41) |
| BRCA2 | 90 | 0.64 (0.49-0.79) | 0.6 (0.46-0.75) | RAD51D | 0.2 (0-0.55) | 0.19 (0-0.52) |
| BRCA2 | 100 | 0.88 (0.79-0.98) | 0.83 (0.7-0.95) | RAD51D | 0.38 (0-0.97) | 0.33 (0-0.86) |
| RAD51C | 1 | 0.06 (0-0.15) | 0.08 (0-0.19) | BRIP1 | 0.03 (0-0.06) | 0.03 (0-0.07) |
| RAD51C | 10 | 0.12 (0-0.29) | 0.14 (0-0.32) | BRIP1 | 0.05 (0-0.11) | 0.06 (0-0.13) |
| RAD51C | 25 | 0.16 (0-0.38) | 0.18 (0-0.41) | BRIP1 | 0.07 (0-0.15) | 0.08 (0-0.17) |
| RAD51C | 50 | 0.23 (0-0.51) | 0.23 (0-0.53) | BRIP1 | 0.1 (0-0.21) | 0.11 (0-0.22) |
| RAD51C | 75 | 0.31 (0-0.67) | 0.3 (0-0.66) | BRIP1 | 0.14 (0-0.29) | 0.14 (0-0.29) |
| RAD51C | 90 | 0.4 (0-0.84) | 0.38 (0-0.8) | BRIP1 | 0.19 (0-0.38) | 0.18 (0-0.36) |
| RAD51C | 100 | 0.66 (0.14-1) | 0.59 (0.08-1) | BRIP1 | 0.36 (0.04-0.68) | 0.32 (0.02-0.61) |
| ATM | 1 | 0.06 (0.03-0.09) | 0.08 (0.04-0.11) | All women | 0.03 (0.02-0.03) | 0.04 (0.03-0.04) |
| ATM | 10 | 0.12 (0.06-0.17) | 0.13 (0.07-0.19) | All women | 0.05 (0.05-0.06) | 0.06 (0.05-0.07) |
| ATM | 25 | 0.16 (0.08-0.23) | 0.17 (0.09-0.25) | All women | 0.07 (0.06-0.08) | 0.08 (0.07-0.09) |
| ATM | 50 | 0.21 (0.12-0.31) | 0.22 (0.13-0.32) | All women | 0.1 (0.09-0.11) | 0.11 (0.09-0.12) |
| ATM | 75 | 0.29 (0.17-0.41) | 0.29 (0.17-0.41) | All women | 0.14 (0.13-0.15) | 0.14 (0.12-0.15) |
| ATM | 90 | 0.38 (0.23-0.52) | 0.36 (0.22-0.5) | All women | 0.19 (0.17-0.21) | 0.18 (0.16-0.19) |
| ATM | 100 | 0.64 (0.46-0.82) | 0.57 (0.39-0.75) | All women | 0.35 (0.32-0.39) | 0.31 (0.27-0.34) |
| PALB2 | 1 | 0.06 (0.01-0.1) | 0.07 (0.01-0.13) |  |  |  |
| PALB2 | 10 | 0.11 (0.02-0.2) | 0.12 (0.02-0.22) |  |  |  |
| PALB2 | 25 | 0.15 (0.03-0.27) | 0.15 (0.03-0.28) |  |  |  |
| PALB2 | 50 | 0.2 (0.05-0.36) | 0.2 (0.04-0.36) |  |  |  |
| PALB2 | 75 | 0.28 (0.07-0.48) | 0.26 (0.07-0.46) |  |  |  |
| PALB2 | 90 | 0.36 (0.11-0.61) | 0.33 (0.09-0.56) |  |  |  |
| PALB2 | 100 | 0.62 (0.3-0.93) | 0.53 (0.22-0.84) |  |  |  |

**Supplementary Table 5:**
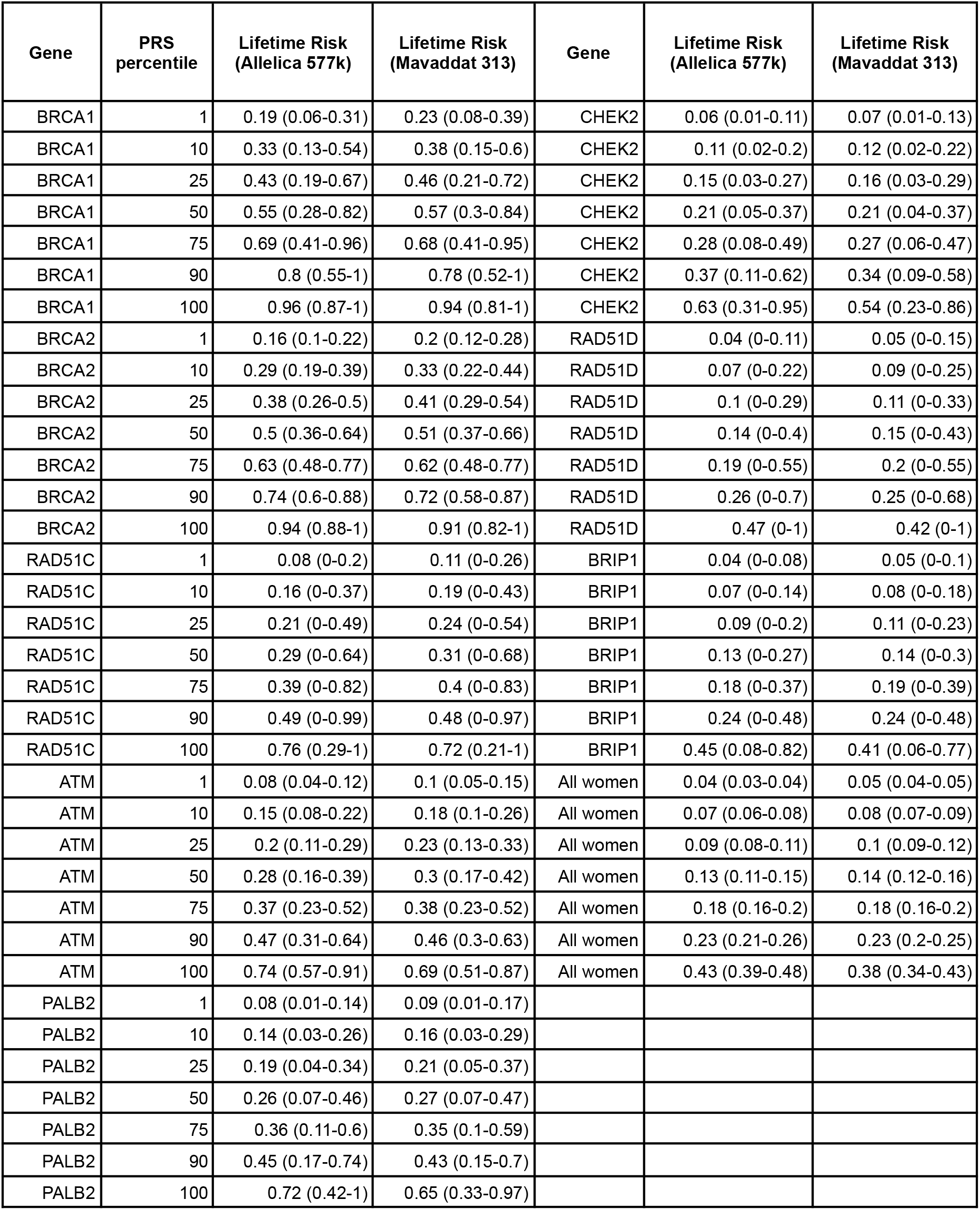
Predicted Lifetime risk (risk by age 79) and 95% confidence intervals for carriers of pathogenic mutations with family history of breast cancer at different PRS strata using the Allelica 577k and Mavaddat 313.

**Supplementary Table 6:**
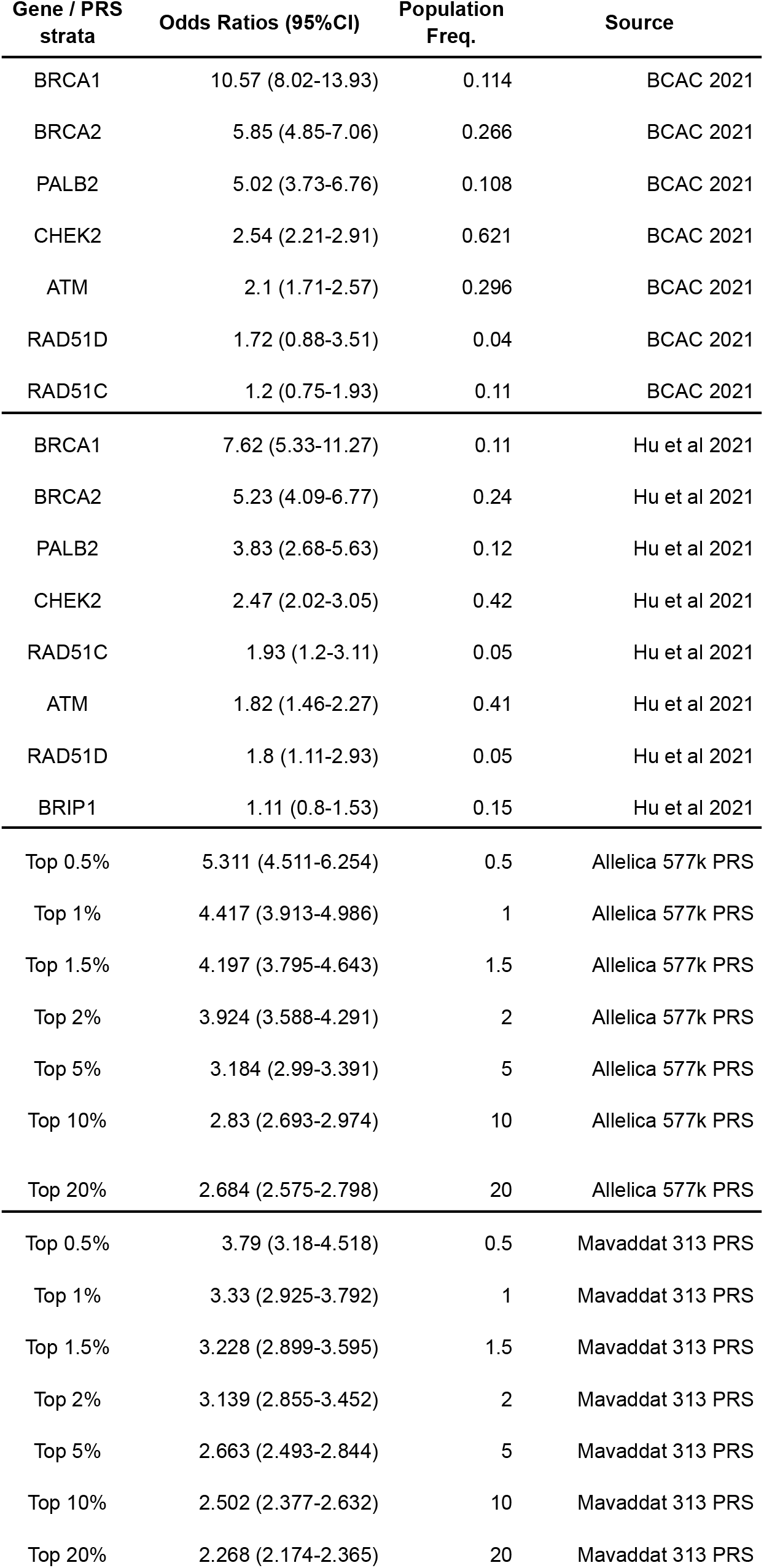
Odds Ratios for carriers of pathogenic mutations in 8 BC susceptibility genes and top strata of the Allelica 577k and Mavaddat 313 SNP PRS.

**Supplementary Figure:**
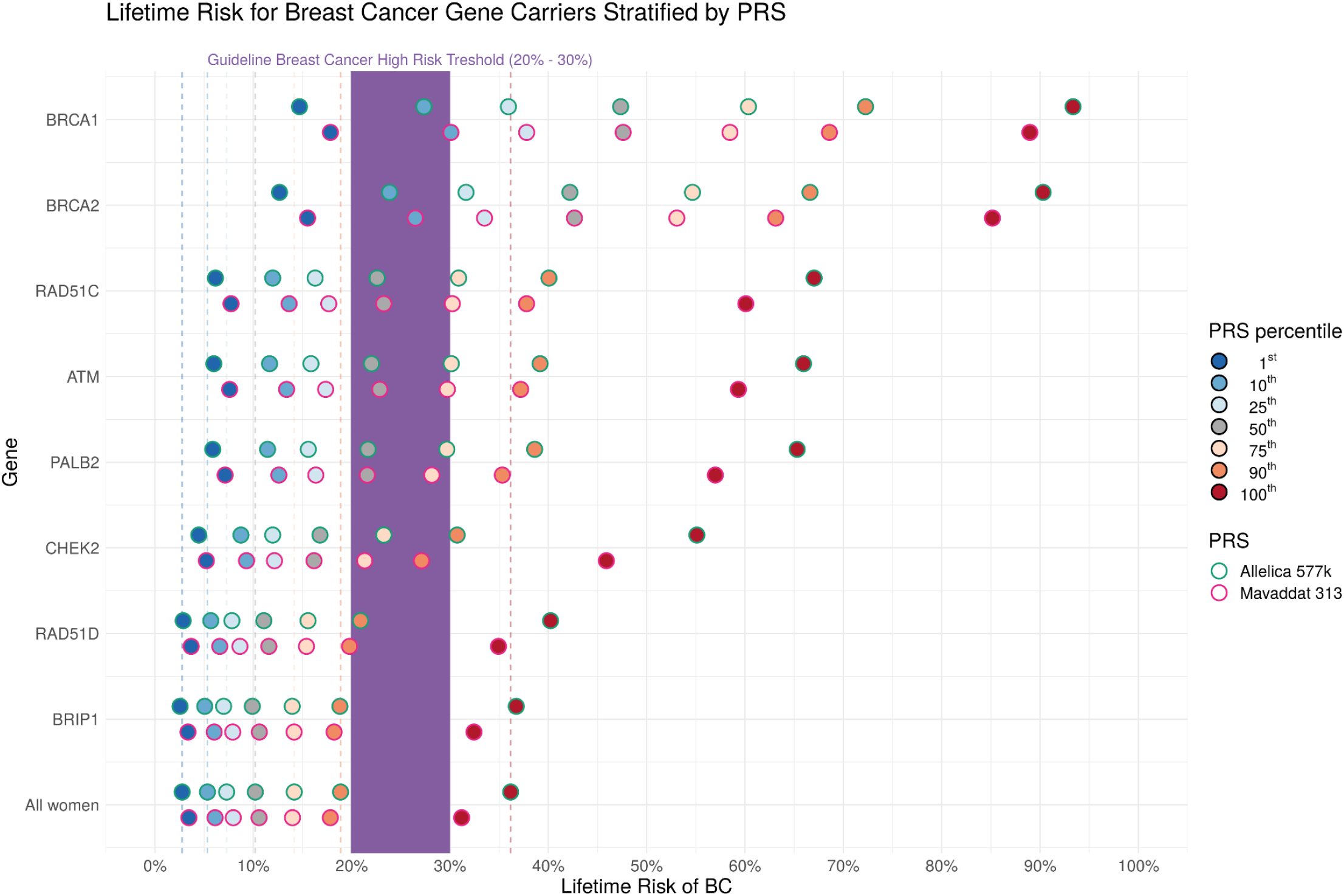
PRS modulates the penetrance of 8 BC susceptibility genes. As Figure 1 with the inclusion of percentiles of the Mavaddat 313 SNP PRS ^17^. We identified carriers of pathogenic variants in 8 breast cancer susceptibility genes in whole exome sequence data in 86,385 women in the UK Biobank. For each gene we show the predicted lifetime risk for carriers across a range of different PRS percentiles for the Allelica 577k PRS. An equivalent model without carrier status was built using all Non carriers (Non carriers). Dashed lines show lifetime risk for percentiles of the Allelica 577k PRS distribution in Non-carriers. American Cancer Association guidelines state that a threshold of 20% lifetime risk equates to High Risk ^20^. We estimated lifetime risk of BC with a Cox proportional hazards regression for each gene separately in a model that included case/control status, age of enrollment and age of disease in the survival function, and carrier status, PRS, 4 PCs and genotyping array as covariates.

## Notes

### Competing Interest Statement

All authors are employees of Allelica Inc. GB, GBB, PD own stock in Allelica Inc.

### Funding Statement

This work was supported by internal reasearch and development funding from Allelica Inc.

### Author Declarations

UK Biobank received ethical approval from the North West Multi-Center Research Ethics Committee (11/NW/03820).

